# Enhanced Insights into Alcohol Use Disorder from Lifestyle, Background, and Family History in a Large-Scale Machine Learning Study

**DOI:** 10.64898/2026.03.01.26347358

**Authors:** Chenlan Wang, Gaojian Huang, Yue Luo, Weihua Zhou

## Abstract

Alcohol Use Disorder (AUD) is a multifactorial condition with severe individual and societal impacts. Extending our 2024 study, this work examines lifestyle, background, and family history determinants of AUD using an expanded dataset from the All of Us Research Program. The updated analysis includes approximately 2.5 times more participants than the prior study, enabling improved statistical power and evaluation of result stability over time. Using interpretable machine learning models and statistical analyses, we identified annual income, residential stability, recreational drug use, sex/gender, marital status, education, and family history as key contributors to AUD risk. Annual income remained the most influential predictor across both datasets, while other feature rankings showed modest shifts. Family history factors continued to demonstrate non-linear effects, with close relatives’ AUD status remaining influential despite differences between statistical association and predictive importance. In predicting AUD versus non-AUD status, Random forest models achieved the highest classification accuracy (81%), consistent with 2024 results but with improved precision for identifying AUD cases. Overall, the findings confirm the robustness of previously identified AUD determinants and underscore the need for coordinated, multi-level prevention strategies addressing behavioral, familial, and structural factors contributing to AUD.

## INTRODUCTION

Alcohol use often begins as a socially accepted behavior associated with relaxation, celebration, or coping with stress. However, sustained and excessive consumption can produce neurobiological and behavioral changes that increase the risk of developing Alcohol Use Disorder (AUD). Repeated alcohol exposure alters neural circuits involved in reward, stress regulation, and executive control (Hanson, Williams, Bangasser, & Peña, 2021), leading to tolerance, physiological dependence, and impaired self-regulation (Elvig et al., 2021; Hull & Slone, 2004). As drinking intensifies, individuals may require increasing amounts of alcohol to achieve desired effects and experience withdrawal symptoms in its absence. These neuroadaptations (Brodie, 2026), combined with psychological vulnerabilities (e.g., stress, trauma, maladaptive coping) and environmental influences (e.g., social norms, availability, family history) (Cavicchioli, Vassena, Movalli, & Maffei, 2018; Chartier, Karriker-Jaffe, Cummings, & Kendler, 2017), shift alcohol use from controlled behavior to compulsive consumption despite negative consequences.

At the population level, these mechanisms translate into a substantial public health and economic burden. In the United States, AUD affects approximately 29.1% of individuals over their lifetime, with 13.9% meeting diagnostic criteria within a given 12-month period (Grant et al., 2015). Beyond health outcomes, excessive alcohol use imposes a considerable economic cost, estimated at about $249 billion annually in the U.S., driven by healthcare expenditures, lost productivity, criminal justice involvement, and other societal impacts (Ahn, 2024). This burden underscores the far-reaching implications of AUD and the urgent need for effective prevention, intervention, and treatment strategies.

Given the prevalence and societal impact of AUD, identifying contributing factors is critical for informing prevention and treatment. Although genetic and environmental influences contribute approximately equally to AUD risk (Pace & Samet, 2016), lifestyle behaviors, background characteristics, and family history represent particularly informative domains for assessment and intervention. Lifestyle factors capture modifiable behaviors such as alcohol use patterns, stress exposure, sleep behaviors, and coping strategies (Cavicchioli et al., 2018). Background characteristics, including socioeconomic status, education, and exposure to trauma, shape vulnerability by influencing access to resources and adaptive coping capacity (Lasserre et al., 2022), while family history reflects both inherited susceptibility and early environmental exposure (Edwards, Sundquist, Sundquist, Kendler, & Larsson Lönn, 2021). Examining these domains enables a comprehensive understanding of how contextual and behavioral factors interact with neurobiological mechanisms in the progression from alcohol use to AUD.

Building on prior work by Wang, Huang, and Luo (2025), the present study extends previous analyses by incorporating additional data, resulting in a sample size approximately twice that reported in 2024 and improved statistical power to detect modest effects. The expanded and more diverse dataset also enables evaluation of the robustness and consistency of identified associations across demographic and contextual subgroups, which is particularly important for multifactorial conditions like AUD where risk arises from complex interactions and model performance may be sensitive to sample composition.

### Objectives

Our prior study used survey data from the All of Us Program’s Registered Tier Dataset (v7) to examine how lifestyle, personal background, and family history affect Alcohol Use Disorder (AUD) risk among 6,016 participants (Wang et al., 2025). Key determinants from 31 factors were identified using decision tree analysis, and machine learning models, including decision trees, random forests, and Naive Bayes, were applied to predict AUD risk, with random forests achieving the highest accuracy at 82%. While the prior study provided valuable insights and predictive models, its findings were constrained by sample size and limited generalizability across demographic subgroups.

The objective of this study is to continue to examine the associations between lifestyle behaviors, individual background, and family history and the risk of AUD using an expanded dataset. By replicating and extending prior analyses from Wang et al. (2025) with approximately twice the sample size reported in 2024, this study aims to assess the robustness and generalizability of previously identified relationships, improve statistical power for detecting modest effects, and evaluate the consistency of contributing factors across demographic and contextual subgroups.

Specifically, the study aims to: 1) Identify the key determinants of AUD to understand how lifestyle, background, and family history influence individual susceptibility to AUD; 2) Develop predictive models of AUD risk based on these determinants, informing targeted prevention and intervention strategies; and 3) Track and analyze temporal changes in these factors, quantifying shifts in key AUD risk determinants and evaluating the consistency of contributing factors across demographic and contextual subgroups over time.

## METHODS

### Data

The data comes from the All of Us Research Program’s Registered Tier Dataset (v8). By 2024, the dataset included 409,420 enrolled individuals, with an additional 224,120 participants added in 2025, bringing the total sample size to 633,540. The dataset covers data from a wide U.S. population, including demographic information, lifestyle and health surveys, electronic health records, and physical measurements. Additional data types, such as wearable device data and genomic information, are available through secure data tiers, supporting comprehensive population health research (The All of Us Research Program Genomics Investigators, 2024). For this AUD study, data were extracted from the All of Us dataset, mainly in Surveys, to capture lifestyle factors (e.g., tobacco, alcohol, and drug use in “Lifestyle”), background characteristics (e.g., demographics and other socioeconomic status in “The Basics”), and family history of alcohol or substance use disorders (in “Personal and Family Health History”). These variables were selected to examine how behavioral, contextual, and familial factors contribute to individual susceptibility to AUD. Details, including the labels and brief description of the survey questions, can be found in Wang et al. (2025). A total of 33 factors (i.e., features) are considered in this study, consistent with Wang et al. (2025).

### Data Preprocess

Each of the three surveys included over 200,000 participants in 2025, roughly double the number from 2024, when each survey had over 100,000 participants. Consistent with the 2024 analysis, only participants who completed all three surveys were included. Responses marked as “Skip,” “Prefer Not To Answer,” “Don’t Know,” or “Response removed due to invalid value,” were considered incomplete and excluded from the analysis. After applying these criteria, the final analytic dataset comprised 15,090 participants with full information. In 2024, this number was 6,016, so the 2025 dataset is roughly 2.5 times larger, reflecting a substantially increased scale compared to last year. In 2025, among the participants, 3,854 (26%) have experienced AUD, while 11,236 (74%) have never had AUD in their lifetime. Compared to 2024, these numbers represent increases of approximately 2.67 times for participants with AUD and 2.46 times for those without AUD. Consistent with 2024, the 2025 data show that sex at birth and gender identity responses were identical, so we excluded gender to avoid redundancy. Likewise, 99.6% of participants reported ever drinking alcohol, similar proportion as in 2024, though the total number of participants is 2.5 times higher, making this variable uninformative for assessing AUD. In total, 31 features were retained for analysis and prediction.

#### Models

This study replicated our previous work (Wang et al., 2025) to a larger, updated dataset. To examine how individual attributes relate to AUD (Objective 1), we analyzed feature importance using decision trees (Kingsford & Salzberg, 2008) due to their natural ability to rank features. Top factors were further examined via data visualization. When visual inspection did not reveal clear relationships, we applied the Chi-Square Test of Independence (McHugh, 2013) for binary nominal variables. Next (Objective 2), we predicted the likelihood of developing AUD using the identified features, comparing Decision Trees, Random Forests (Breiman, 2001), and Naive Bayes (Rish et al., 2001) for computational efficiency and interpretability. Detailed methodology is provided in Wang et al. (2025). Finally (Objective 3), we examined how feature importance and relationships changed over time with the updated dataset.

All analyses were conducted within the All of Us Research Workbench (Python 3.10.16) to ensure data security, which means no data were downloaded to local workstations. Consistent with our previous study, 80% of the data were used for training and 20% for testing. Both standard holdout validation (Lachenbruch & Mickey, 1968) and 5-fold cross-validation (Stone, 1974) were applied to mitigate the risk of overfitting.

## RESULTS AND DISCUSSION

### Importance of Each Attribute

Figure 1 shows the relative importance of participants’ lifestyle, background, and family history features for AUD from the large-scale dataset. Compared with the 2024 results from Wang et al. (2025) (Table 1), individual annual income (0.0776) remains the most influential factor and continues to be a key determinant of AUD. Years living at the current address (0.0743) has increased in importance, rising from fifth to second place. Recreational drug use (0.0689) is now ranked third, compared to second in 2024. Family history of AUD remains a significant contributor, with AUD-Son, AUD-Sibling, and AUD-Mother continuing to appear among the top 10 factors; AUD-Grandparent is newly included, replacing Marriage. Household size shows comparable importance across years (9th in 2025 vs. 10th in 2024), while education emerges as a new top factor in the 2025 analysis.

**Table 1.** Top 10 factors based on the importance ranking using a decision tree for the 2024 dataset (v7) and the 2025 dataset(v8)

| Factor ranking | 2024 dataset (v7) | 2025 dataset (v8) |
| --- | --- | --- |
| 1 | Income | Income |
| 2 | RecreationDrug | LivingYear |
| 3 | AUD-Grandparent | RecreationDrug |
| 4 | Sex | Sex |
| 5 | LivingYear | AUD-Mother |
| 6 | AUD-Sibling | AUD-Son |
| 7 | AUD-Mother | AUD-Sibling |
| 8 | AUD-Son | AUD-Grandparent |
| 9 | Marriage | HouseholdNum |
| 10 | HouseholdNum | Education |

**Figure 1.**
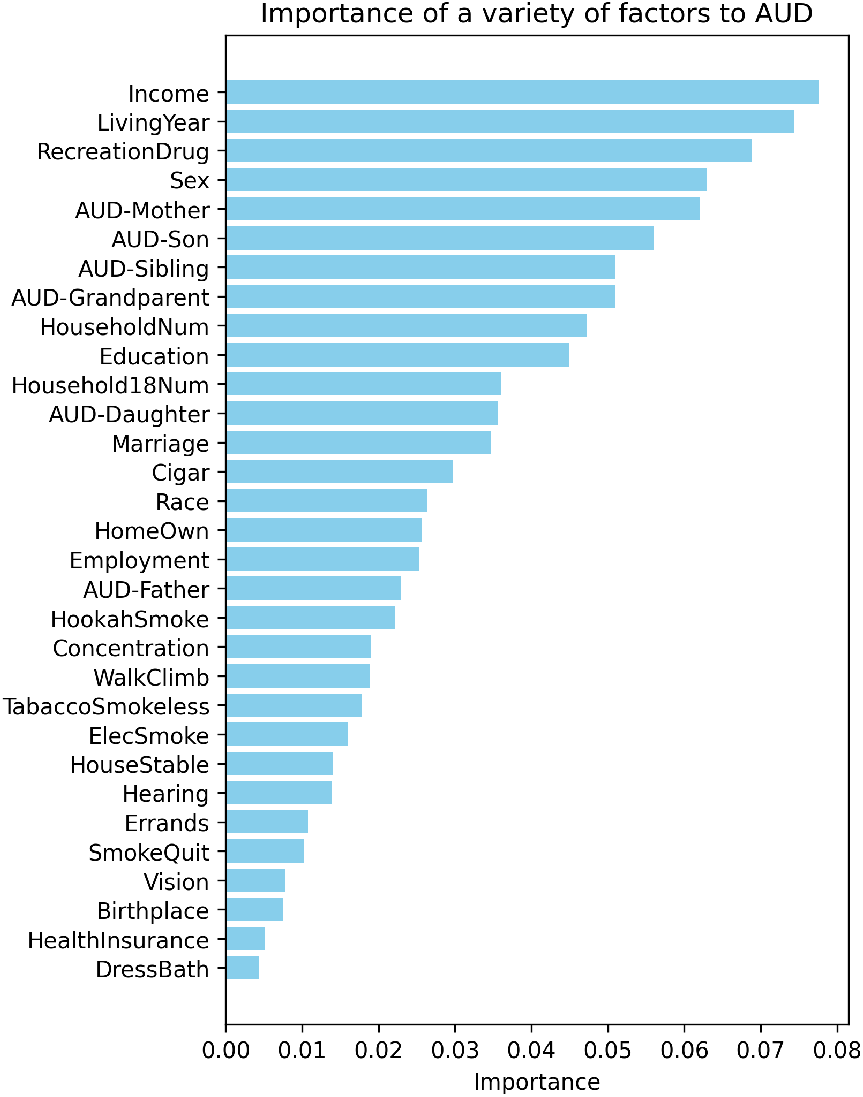
The relative importance of factors from the decision tree in predicting whether an individual has AUD.

### Intervention Opportunities Based on Modifiable Factors

Consistent with our previous work, since sex at birth is fixed, we focused on the ten other top factors, such as family history and behavioral patterns, from *The Basics* and *Personal and Family Health History* surveys to identify modifiable areas for interventions.

Again, we find that family history does not influence AUD risk in a simple linear manner (i.e, does not show a linear relationship with AUD); however, having close relatives with AUD, particularly a mother, sibling, son, or grandparent, remains an important and complex predictor (i.e., such factors listed in Table 1). This pattern suggests that the contribution of family history to AUD risk is non-linear and likely mediated by interactions involving environmental exposures, familial drinking norms, individual adaptive responses, and other factors. Figure 2 further illustrates the importance of each of the six family history factors identified from Figure 1 related to AUD, indicating that AUD in close family members, especially mothers (22.3%), sons (20.1%), grandparents (18.3%), and siblings (18.3%), has the strongest influence on AUD risk.

**Figure 2.**
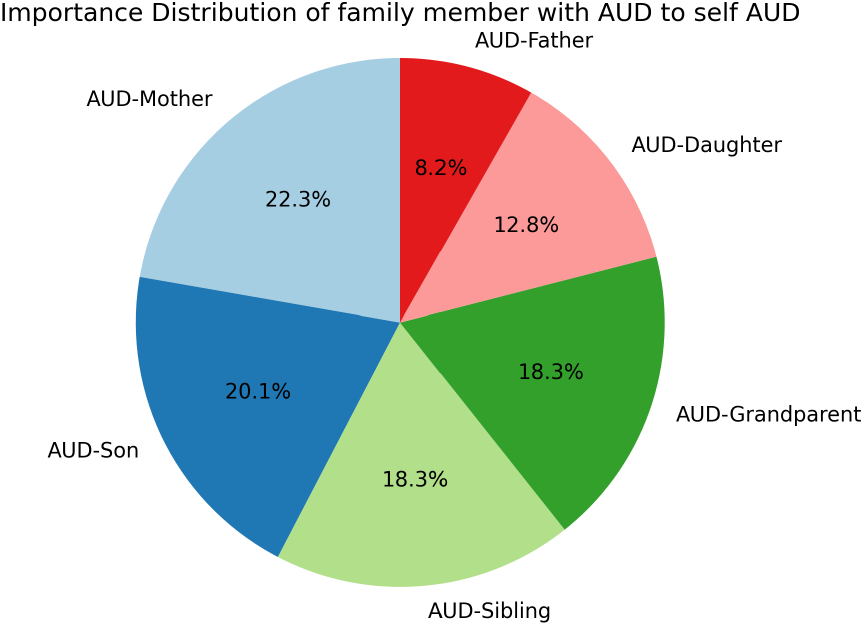
The relative importance of family members’ AUD status in predicting individual AUD.

**Figure 3.**
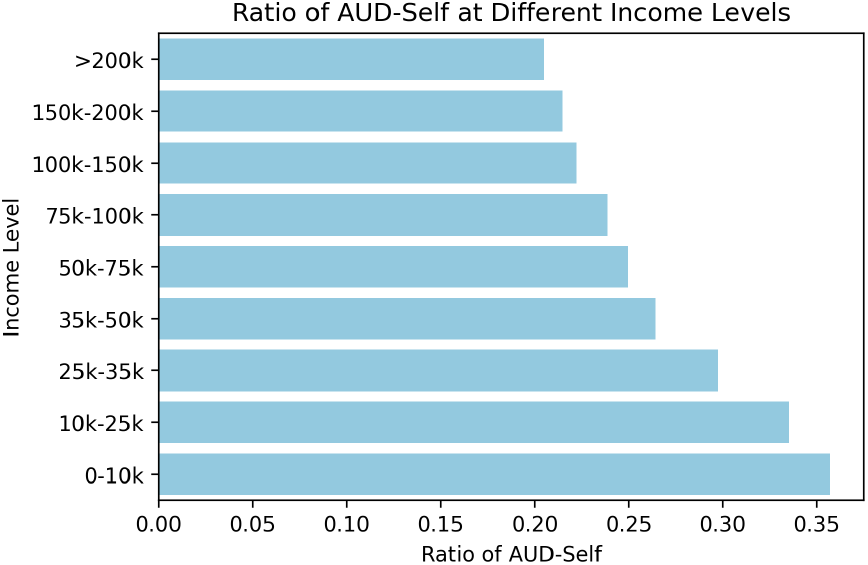
At each annual income level, the ratio of participants with AUD to the total number of participants at that income level is displayed.

**Figure 4.**
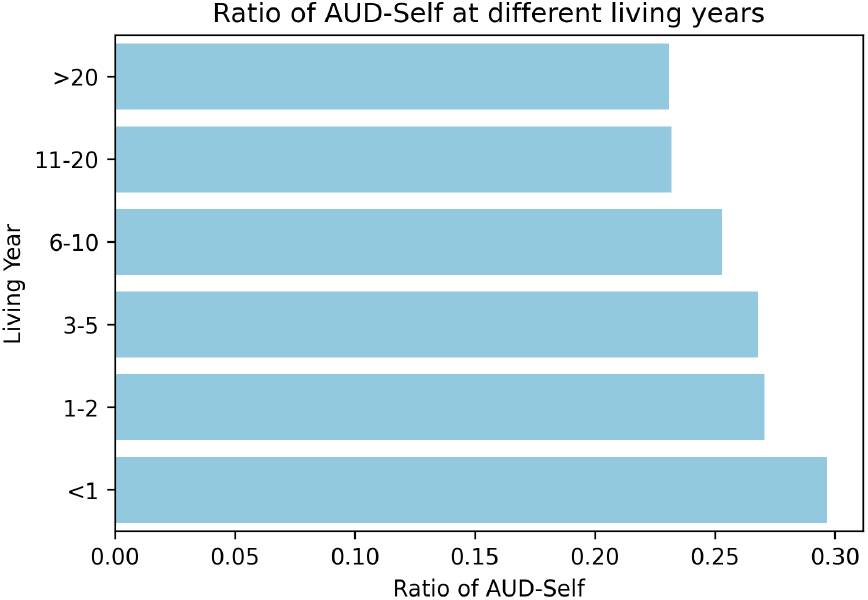
Given the duration of residence at the current address, the ratio of participants with AUD to the total number of participants sharing the same length of residence.

**Figure 5.**
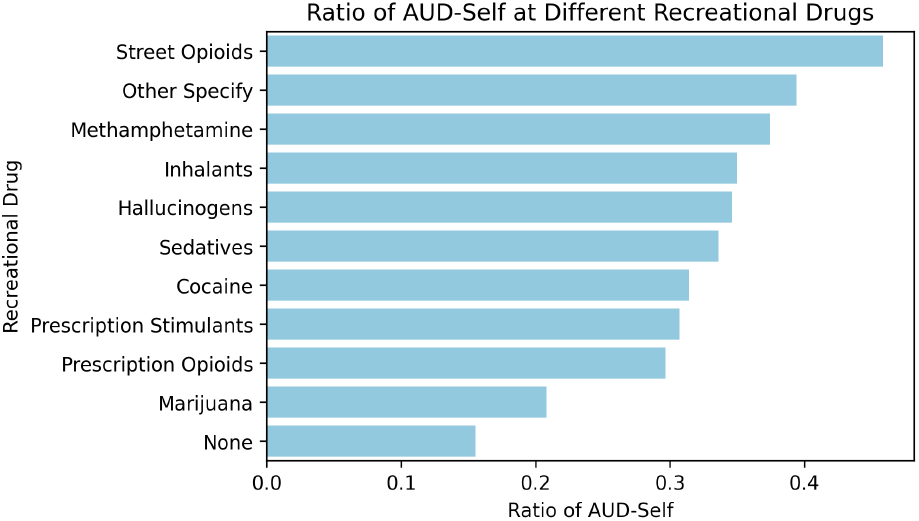
For each type of recreational drug, the ratio of participants with AUD to the total number of participants who have used that drug is presented.

**Figure 6.**
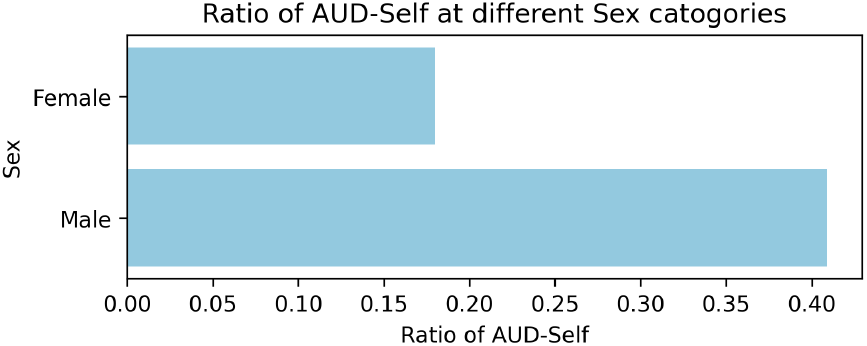
The ratio of participants with alcohol use disorder to the total number of participants, categorized by sex at birth, is presented.

**Figure 7.**
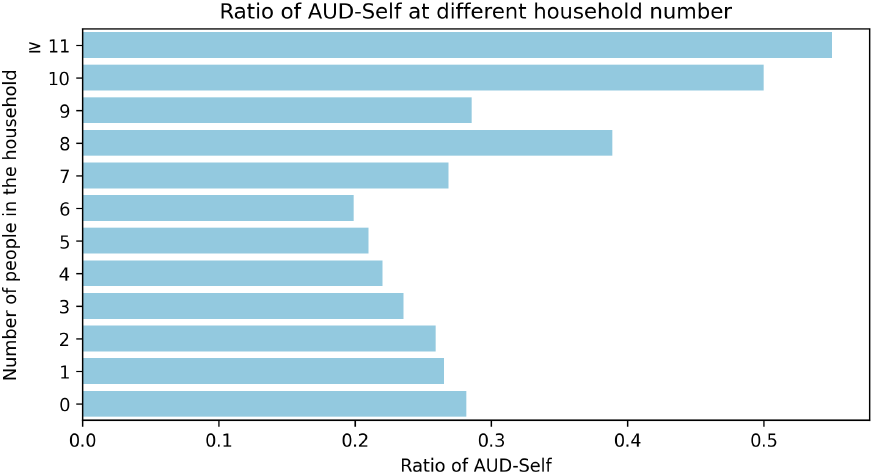
The ratio of participants with alcohol use disorder to the total number of participants, categorized by household number, is presented.

**Figure 8.**
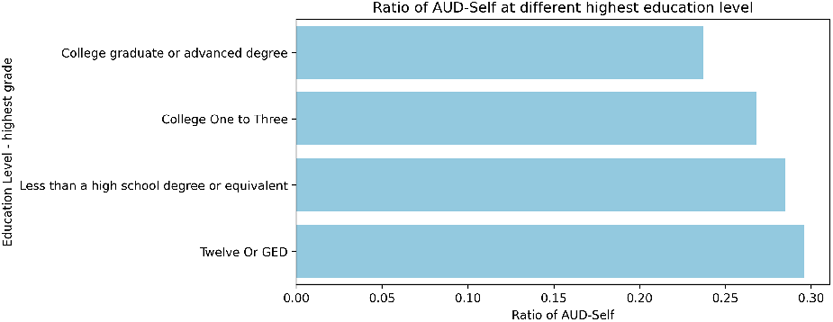
The ratio of participants with alcohol use disorder to the total number of participants is presented, categorized by highest education level.

**Figure 9.**
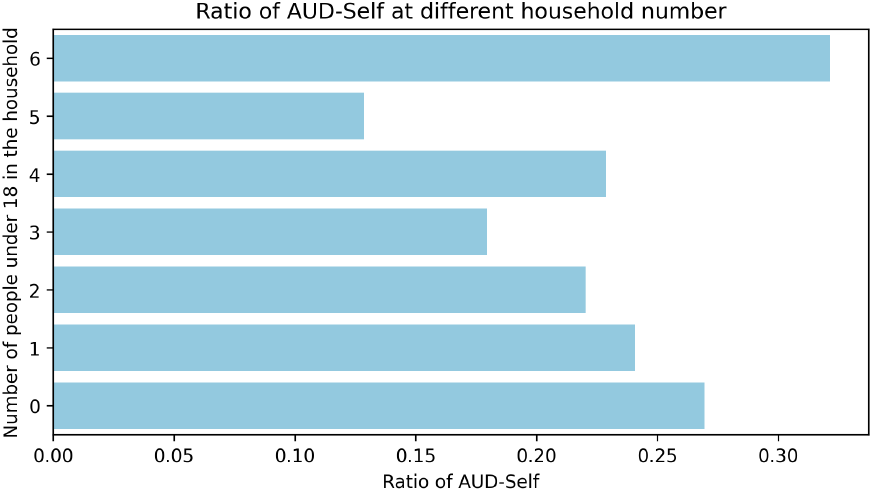
The ratio of participants with alcohol use disorder to the total number of participants, categorized by Household Number of Individuals Under 18, is presented.

**Figure 10.**
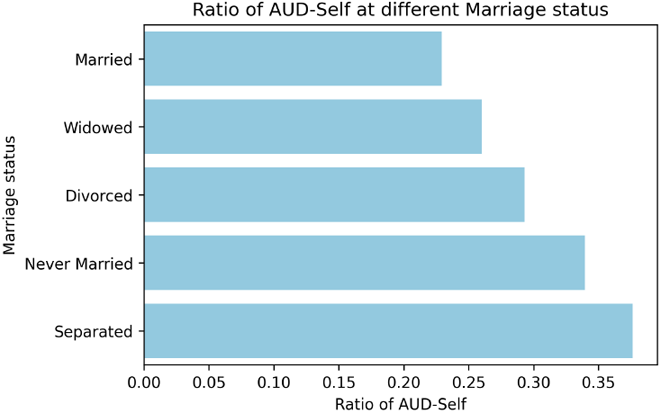
The ratio of participants with alcohol use disorder to the total number of participants, categorized by marriage status, is presented.

Alternatively, using the Chi-Square Test of Independence, we find that three of the six AUD family factors are significantly associated with AUD this year (p < 0.001), compared with four of six in 2024.

- AUD-Grandparent: χ^2^ = 42.80, *p* = 6.06 × 10^−11^
- AUD-Sibling: χ^2^ = 42.35, *p* = 7.64 × 10^−11^
- AUD-Father: χ^2^ = 99.10, *p* = 2.40 × 10^−23^

The results indicate that AUD-Father has the strongest association with AUD-Self, followed by AUD-Grandparent and AUD-Sibling, the same rank as in 2024. This reflects statistical association but not necessarily classification importance.

The discrepancy between the decision tree feature importance and chi-square results reflects fundamental differences in the analytical approaches. Chi-square tests evaluate marginal statistical associations between individual variables and AUD, identifying whether a factor is independently associated with the outcome. In contrast, decision tree models assess predictive importance within a multivariate, non-linear framework, where variables may contribute through interactions with other factors or through hierarchical decision rules. As a result, family history variables that are not statistically significant in chi-square analyses may still play a meaningful role in classification performance when considered with other predictors.

Next, we find the other top factors from *The Basics* Survey all exhibit a comparably strong linear relationship with AUD.

#### Annual Income

Lower annual income is associated with an increased risk of AUD relative to higher income levels. For example, 35.7% of participants earning less than $10, 000 have experienced AUD, up from 31.7% in 2024. In contrast, 20.5% of those earning over $200, 000 have experienced AUD, slightly higher than 19.6% in 2024. With the total participants now 2.5 times larger, the absolute increases in number are even greater.

#### Living Years

Overall, AUD ratios range from 23.1% to 29.7%, compared with 21.7% to 26.2% in 2024. Shorter duration of residence (i.e., less than one year) at the current address is associated with increased AUD prevalence (29.7%, up from 26.2% in 2024). This may reflect stress from transitions, lack of support networks, or use of alcohol as a coping strategy. Unlike in 2024, a clear linear trend now emerges: the shorter the residence, the higher the likelihood of developing AUD.

#### Recreational Drug Use

Individuals without a history of recreational drug use demonstrate the lowest prevalence of AUD. Specifically, 15.5% of non-users have AUD (up from 15.0% in 2024), compared with 45.9% of those who have used street opioids (down from 51.2% in 2024).

#### Sex

AUD prevalence is lower among females than males. Specifically, 18.0% of participants with AUD are female (16.1% in 2024), while 40.9% are male (38.2% in 2024). Overall, the total number of participants with AUD is increasing for both genders.

#### Household Number

Individuals residing in households with six members exhibit the lowest AUD-Self ratio at 19.9%, whereas those in households with more than eleven members demonstrate the highest AUD-Self ratio at 55%. This indicates a marked increase in the prevalence of AUD-Self as household size increases. These findings highlight the potential influence of household size on the risk or reporting of AUD. Larger households may present unique social, economic, or environmental factors that contribute to higher AUD prevalence, or they may increase the likelihood of observation and self-reporting of AUD symptoms.

#### Education Level

Educational attainment below the college level is associated with a higher prevalence of AUD. Specifically, 23.7% of participants with AUD have completed a college degree or higher (22.3% in 2024), while 29.6% report a highest level of education of high school or general educational development (27.5% in 2024), highlighting the potential impact of education on AUD risk, though it does not imply a causal relationship.

#### Household Number of Individuals Under 18

Individuals residing in households with five members under the age of 18 report the lowest AUD-Self ratio at 12.85%. In contrast, households with more than six members under 18 exhibit the highest AUD-Self ratio at 32.14%. This pattern reveals a substantial increase in the prevalence of AUD-Self as the number of younger household members rises. These findings underscore the potential impact of household size under 18 on both the risk and reporting of AUD. Larger households with more minors may encounter distinct social, economic, or environmental factors that elevate the risk of AUD or enhance the likelihood of identifying and self-reporting AUD symptoms.

#### Marriage Status

Marital status is associated with AUD prevalence, with never-married and separated individuals exhibiting greater risk relative to married or widowed individuals. Specifically, 22.9% of participants with AUD are married (20.9% in 2024), while 37.6% are separated (34.5% in 2024), suggesting that relationship stability and support may help reduce AUD risk.

#### Marriage status

### Prediction: AUD or NonAUD

Table 2 and Table 3 summarizes the classification performance of AUD verses nonAUD via three modeling approaches: Decision Tree, Random Forests, and Naive Bayes. Among the three machine learning techniques, Random Forests perform best overall, achieving 81% accuracy (same as in 2024). The model performs well in identifying non-AUD cases, and when it predicts AUD, it is generally accurate, with a precision of 80%, more accurate than in 2024 (75%). Notably, the precision for AUD has increased across all three models. However, recall for AUD is low (32%), meaning many actual AUD cases are missed. This imbalance likely reflects the data distribution (767 with AUD vs. 2,251 without AUD, more than 2 times the numbers in 2024). Addressing class imbalance through methods such as resampling or stratified cross-validation could improve the detection of AUD cases in future work.

**Table 2.** Classification accuracy when predicting AUD vs. NonAUD. Accuracy-1 fold represents results from a holdout validation, whereas Accuracy-5 fold corresponds to 5-fold cross-validation.

| ML Technique | Decision Trees | Random Forests | Naive Bayes |
| --- | --- | --- | --- |
| Accuracy-1 fold | 0.72 | 0.81 | 0.70 |
| Accuracy-5 fold | 0.71 | 0.81 | 0.70 |

**Table 3.** Classification performance metrics when predicting AUD vs. NonAUD using holdout validation.

| ML Technique | AUD-Self | precision | Recall | $f_1$ -score |
| --- | --- | --- | --- | --- |
| Decision Trees | no | 0.83 | 0.79 | 0.81 |
|  | yes | 0.46 | 0.51 | 0.48 |
| Random Forests | no | 0.81 | 0.97 | 0.88 |
|  | yes | 0.80 | 0.32 | 0.46 |
| Naive Bayes | no | 0.80 | 0.80 | 0.80 |
|  | yes | 0.41 | 0.40 | 0.40 |

### Interpretation from a Human Factors Perspective

Our study highlights the multifactorial nature of AUD risk. Interaction of Individual and Structural Determinants: Income, education, and marital stability demonstrate how individual behaviors cannot be disentangled from structural environments. Low-income participants not only face greater financial stress but also reduced access to healthcare and supportive networks, creating reinforcing cycles that exacerbate harmful drinking. From a systems design perspective, interventions targeting AUD may extend beyond the clinic to address socioeconomic inequities.

Family and Social Contexts as Risk Pathways: Family history of AUD remains a robust predictor, but its effect is mediated through environment, upbringing, and coping styles. The fact that paternal and sibling AUD showed stronger associations than maternal factors may reflect gendered differences in modeling behaviors or patterns of household dynamics. Moreover, household size findings suggest that living arrangements, whether crowded or isolated, shape exposure and stress in complex ways. Human factors research could explore how these relational environments either buffer or amplify vulnerabilities. Occupational and Policy Implications: The predictive models illustrate how risk factors cluster, suggesting that interventions could be tailored not only to individuals but also to groups with shared profiles, such as those with unstable housing, low income, or comorbid substance use. From a human factors’ perspective, workplaces represent critical sites of intervention. At the policy level, regulating alcohol availability and marketing may mitigate the population-wide amplification of risk identified in this study.

Systems-Level Recommendations: For system-level recommendations, Human factors insights argue for a multi-level prevention strategy: (1) Individual level: Screening and early interventions for those with family AUD history or psychiatric comorbidities; (2) Community level: Strengthening social networks to mitigate risks associated with relocation and unstable households; (3) Policy level: Expanding economic support for vulnerable populations; (4) Workplace level: Implementing stress-reduction programs in high-risk professions. Such approaches reframe AUD not as an individual failure but as a predictable outcome of systemic pressures that can be redesigned.

## CONCLUSION

Using interpretable machine learning and statistical analyses, this study identified income, recreational drug use, residential stability, sex/gender, marital status, education, and family history as key factors associated with Alcohol Use Disorder (AUD). Annual income remained the most influential predictor across both the current and 2024 datasets, while other feature rankings showed moderate variation over time. Chi-square analyses revealed consistent patterns of association, and among three predictive models trained on lifestyle, background, and family history variables, random forests achieved the highest accuracy (81%). Overall, the findings emphasize that AUD risk is shaped by interacting social, behavioral, and familial factors, highlighting the need for coordinated, multi-level prevention strategies rather than isolated interventions.

## Data Availability

All data produced are available online at the NIH AllofUs research workbench titled "Assessing Alcohol Use Disorder using Machine Learning Techniques-updated with v8".

## ACKNOWLEDGEMENT

This work was made possible by the contributions of participants in the All of Us Research Program. We further acknowledge the National Institutes of Health for granting access to the data resources utilized in this study.

## REFERENCES

Ahn, H. (2024). Three essays in population health, health economics and outcomes research. The Pennsylvania State University.

Breiman, L. (2001). Random forests. Machine learning, 45, 5–32.

Brodie, M. S. (2026). Elucidating noradrenergic neuroadaptations of the central amygdala in alcohol use disorder. Biological Psychiatry, 99(1), 2–3.

Cavicchioli, M., Vassena, G., Movalli, M., & Maffei, C. (2018). Addictive behaviors in alcohol use disorder: dysregulation of reward processing systems and maladaptive coping strategies. Journal of addictive diseases, 37(3-4), 173–184.

Chartier, K. G., Karriker-Jaffe, K. J., Cummings, C. R., & Kendler, K. S. (2017). Environmental influences on alcohol use: Informing research on the joint effects of genes and the environment in diverse us populations. The American journal on addictions, 26(5), 446–460.

Edwards, A. C., Sundquist, K., Sundquist, J., Kendler, K. S., & Larsson Lönn, S. (2021). Genetic and environmental influences on the progression from alcohol use disorder to alcohol-related medical conditions. Alcoholism: Clinical and Experimental Research, 45(12), 2528–2535.

Elvig, S. K., McGinn, M. A., Smith, C., Arends, M. A., Koob, G. F., & Vendruscolo, L. F. (2021). Tolerance to alcohol: A critical yet understudied factor in alcohol addiction. Pharmacology Biochemistry and Behavior, 204, 173155.

Grant, B. F., Goldstein, R. B., Saha, T. D., Chou, S. P., Jung, J., Zhang, H., … others (2015). Epidemiology of dsm-5 alcohol use disorder: results from the national epidemiologic survey on alcohol and related conditions iii. JAMA psychiatry, 72(8), 757–766.

Hanson, J. L., Williams, A. V., Bangasser, D. A., & Peña, C. J. (2021). Impact of early life stress on reward circuit function and regulation. Frontiers in Psychiatry, 12, 744690.

Hull, J. G., & Slone, L. B. (2004). Alcohol and self-regulation. Handbook of self-regulation, 466–491.

Kingsford, C., & Salzberg, S. L. (2008). What are decision trees? Nature biotechnology, 26(9), 1011–1013.

Lachenbruch, P. A., & Mickey, M. R. (1968). Estimation of error rates in discriminant analysis. Technometrics, 10(1), 1–11.

Lasserre, A. M., Imtiaz, S., Roerecke, M., Heilig, M., Probst, C., & Rehm, J. (2022). Socioeconomic status, alcohol use disorders, and depression: a population-based study. Journal of affective disorders, 301, 331–336.

McHugh, M. L. (2013). The chi-square test of independence. Biochemia medica, 23(2), 143–149.

Pace, C. A., & Samet, J. H. (2016). Substance use disorders. Annals of internal medicine, 164(7), ITC49–ITC64.

Rish, I., et al. (2001). An empirical study of the naive bayes classifier. In Ijcai 2001 workshop on empirical methods in artificial intelligence (Vol. 3, pp. 41–46).

Stone, M. (1974). Cross-validatory choice and assessment of statistical predictions. Journal of the royal statistical society: Series B (Methodological), 36(2), 111–133.

The All of Us Research Program Genomics Investigators. (2024). Genomic data in the all of us research program. Nature, 627, 340–346. doi: 10.1038/s41586-023-06957-x

Wang, C., Huang, G., & Luo, Y. (2025). Assessing alcohol use disorder: Insights from lifestyle, background, and family history with machine learning techniques. In Proceedings of the international symposium on human factors and ergonomics in health care (Vol. 14, pp. 16–20).

